# A comparative analysis of the prevalence of suicidal ideation among depressed and non-depressed pregnant women in rural Bangladesh

**DOI:** 10.1101/2024.07.15.24310425

**Authors:** Rifa Tamanna Mumu, Md Parvez Shaikh, Dipak Kumar Mitra

## Abstract

**Background:** More than 300 million people all over the world succumbed to depressive disorders in 2015. 680 per 100,000 expectant mothers worldwide bear suicidal ideation during the antenatal period. Despite suicidal ideation being a consequence of antenatal depression, there is a scarcity of information on the prevalence of suicidal ideation in depressed and non-depressed pregnant women in rural Bangladesh.

**Objective:** This study is directed to evaluate the point prevalence of suicidal ideation and compare the prevalence between depressed and non-depressed pregnant women in rural Bangladesh.

**Method:** A cross-sectional study was performed in Lohagara, a rural subdistrict in Bangladesh between January 08 and 17, 2024. 351 pregnant women of various trimesters were recruited for the study. The Bengali-translated version of the Edinburgh Postnatal Depression Scale (EPDS) and another structured questionnaire were used for data collection. Data analyses were done by STATA version 17.

**Result:** The point prevalence of suicidal ideation is 11.4% (95% CI: 8.5% to 15.2%). It reveals a similarity between depressed and non-depressed pregnant women. The prevalence of suicidal thoughts in antenatally depressed women accounts for 10.2% (95% CI: 6.1% to 16.6%) and in non-depressed pregnant women it is 12.2% (95% CI: 8.4% to 17.3%) -obtained after analysis.

**Conclusion:** The considerable prevalence of suicidal thoughts among depressed and non-depressed rural pregnant women in Bangladesh underscores the necessity of ensuring additional counseling, care, and support to expectant mothers during their antepartum.

## 1. Introduction

Among the common causes of maternal death in some countries, suicide is one, and suicidal ideation aggravates during pregnancy [1]. 680 in every 100,000 women attempt suicide during their antenatal period globally [2]. A higher prevalence of suicide is observed among Southeast Asian pregnant women (2·19%, 95%CI:1·04–3·68)[3]. In 1.6 to 4.5 per 100,000 live births, women in antenatal and postpartum periods commit suicide in the United States [4]. 20% of postpartum deaths are occurred by suicide, which is regarded as the second most common cause of death among the depressed populace [5]. Anxiety, major depression, and unplanned pregnancy influence the augmentation of suicidal ideation during pregnancy [6].

Approximately, 322 million people all over the world were grappling with depressive disorders in 2015, according to the World Health Organization (WHO) and 27% of them resided in the Southeast Asian territory [7]. Generally, females are more prone to develop mental health disorders like depression than their male counterparts [8]. Certain hormonal changes take part in the antenatal period which enhances the chance of developing depression and raises the percentage in pregnant women than in non-pregnant [9]. The global range of antenatal depression is evaluated as 15% to 65% [10]. The prevalence varies from 5% to 30% [11-13] in high-income countries, while the range is 15.6% to 31.1% in low-income countries [14-16].

The rate of depression in prenatal women is 39% (38.86%, 33.9%-44%) in rural Bangladesh [17] and 14% of depressed women manifest thoughts of self-injury. An unsupportive husband or mother-in-law, family violence, duress due to the family preference for a male child [18], duration of pregnancy, suffering from a previous disease, unintentional pregnancy, having polygamous husbands, and intimate partner violence catalyst the development of depression in the prenatal period [17]. Pregnancy within the first year of the previous one, the inadequacy of social support from allies, and poor health status are conducive to the augmentation of suicidal ideation in adolescent pregnant women in Bangladesh [19].

Though antenatal depression is revealed to have an association with pregnancy-related suicidal ideation in many studies [20, 21], there is no study available in Bangladesh investigating the prevalence of suicidal thoughts in depressed and non-depressed pregnant women. This study is directed to investigate the point prevalence of suicidal ideation and compare the prevalence in depressed and non-depressed women in any trimester of pregnancy in a rural subdistrict in Bangladesh. The notion of the percentage of suicidal ideation in both subgroups will help formulate specific plans and protective strategies to keep expectant mothers’ mental health well and safeguard them from making dreadful decisions.

## 2. Method

### 2.1 Study Design and Setting

A cross-sectional study was performed between January 08 and 17, 2024, in Upazila Health Complex, Lohagara, a government hospital, and Khan General Hospital, Lahuria, a private hospital in Lohagara, a rural sub-district in Narail, situated in the southern part of Bangladesh.

### 2.2 Study Participants

The target population was pregnant mothers of any trimester in the Lohagara, and the sample population was pregnant mothers of any trimesters attending the ANC Corner of Upazila Health Complex, Lohagara, Narail, and Khan General Hospital, Lahuria for antenatal checkups.

### 2.3 Sample Size and Sampling Technique

Considering the prevalence of thoughts of self-harm among pregnant women (*P*_1_) in rural Bangladesh is 14% according to a study conducted in a rural sub-district in Matlab [18], the estimated prevalence of suicidal ideation among non-depressed pregnant women (*P*_2_) is 9%, with a 95% confidence interval and 80% power, 351 data were collected by systematic sampling to minimize bias. Every third patient attending antenatal checkups in both government and private hospitals was selected as a participant in the interview for data collection.

### 2.4 Data Collection Tools

The presence of depression and suicidal ideation was assessed by the Bengali-translated version of the Edinburgh Postnatal Depression Scale (EPDS). This questionnaire consists of 10 questions scoring from 0 to 30. A score of 10 or higher on the assessment indicates probable antenatal depression, and the Item-10 of this questionnaire indicates suicidal ideation. Another structured questionnaire which was also translated into Bengali was used to collect the sociodemographic data of patients. Questionnaires were tested on some target population rather than the study participants and the necessary changes were made before the data collection.

### 2.5 Data Management & Analysis Plan

The data for the study was analyzed by STATA version 17. A chi-squared test was performed to assess the association between depression and non-depression with suicidal ideation. The obtained p-value was used to determine the association between variables.

### 2.6 Ethical Considerations

Ethical permission was obtained from the Institutional Ethics Committee of North South University before data collection (2023/OR-NSU/IRB/1224). Permission letters from Upazila Health Complex, Lohagara, and Khan General Hospital, Lahuria were taken as well. Informed written consents were obtained from pregnant women of 18 years before data collection. Legal guardians of participants signed the written consent form if the participants were under 18. Respondents were assured of the confidentiality of information as well as informed about the purpose, benefits, and potential risks of the study.

## 3. Result

### 3.1 Socio-demographic Characteristics of Pregnant Mothers

This research was conducted with the participation of 351 women in any trimester of gestation. The response rate was 98.5%. The median Edinburgh Postnatal Depression Scale (EPDS) score was 8, with an interquartile range of 4 to 12. The participants had a mean age of 23 years, with an interquartile range of 20 to 27 years. Among the respondents, 5.7% (20) were under 18 years old, while a greater percentage (83.7%) were between the age bracket of 18 to 30 years. The rest 10.6% encompassed women of the thirty-plus age range.

26 participants (7.4%) did not pass the primary school. All respondents were married, and the larger portion were housewives. Only (4%) were employed. 26% of women (91) had a family income of less than 10,000 BDT per month, 58% (203) families earned between 10,000 to 20,000 BDT, and the remaining 16% (56) had a higher monthly income exceeding 20,000 tk.

From a religious perspective, the respondents belonged to two communities; 96.3% (337) of the women were practicing Muslim, and 3.71% (13) were Hindu. **Error! Reference source not found**.

### 3.2 Prevalence of depression among pregnant women

In the study, the scores from the 10 questions of the Edinburgh Postnatal Depression Scale (EPDS) were summed up, creating a new variable ranging from 0 to 30. A cut-off score of 10 or more was used to indicate the presence of depression. The results showed that 137 women (39.03%, with a 95% confidence interval ranging from 34% to 44%) exhibited symptoms of depression during pregnancy and 214 (60.97%, 55.7%-66%) of women didn’t have antenatal depression. **Table 2**.

**Table 1.** Sociodemographic characteristics of pregnant women attending antenatal checkups in Upazila Health Complex, Lohagara, Narail, 2024.

| Sociodemographic status of pregnant women |  |  |
| --- | --- | --- |
| Variables | n | % |
| **Age** |  |  |
| <18 years | 20 | 5.71 |
| 18-30 years | 293 | 83.71 |
| >30 years | 37 | 10.57 |
| **Education** |  |  |
| Below class 5 | 26 | 7.43 |
| Higher | 324 | 92.57 |
| **Employment status** |  |  |
| Employed | 14 | 4 |
| Unemployed | 336 | 96 |
| **Family income (BDT)** |  |  |
| <10,000 | 91 | 26 |
| 10,000-20,000 | 203 | 58 |
| >20,000 | 56 | 16 |
| **Religion** |  |  |
| Hindu | 13 | 3.7 |
| Muslim | 337 | 96.3 |
| Others | 0 | 0 |

**Table 2.** Prevalence of Antenatal Depression in Lohagara, Narail, 2024.

| Prevalence of antenatal depression |  |  |  |
| --- | --- | --- | --- |
| Depression | EPDS score | n | % |
| Yes | 10-30 | 137 | 39.03 |
| No | 0-9 | 214 | 60.97 |

### 3.3 Prevalence of suicidal ideation in pregnancy

A total of 40 women among the 351 had suicidal thoughts during pregnancy which made the overall prevalence of suicidal ideation 11.4% (95% CI: 8.5% to 15.2%). The determined prevalence of suicidal ideation among women suffering from antenatal depression was 10.2% (95% CI: 6.1% to 16.6%). On the contrary, 12.2% (95% CI: 8.4% to 17.3%) of women without prenatal depression had suicidal thoughts. **Table 3**.

**Table 3.** Prevalence of suicidal ideation among depressed and non-depressed pregnant women in Lohagara, Narail, 2024.

| Suicidal Ideation | Antenatal Depression |  | Total |
| --- | --- | --- | --- |
|  | Yes | No |  |
| Yes | 14<br>10.22% | 26<br>12.15% | 40<br>11.40% |
| No | 123<br>89.78% | 188<br>87.85% | 311<br>88.6% |
| **Total** | 137<br>100% | 214<br>100% | 351<br>100% |

## 4. Discussion

This study is directed to assess the prevalence of suicidal ideation among depressed and non-depressed pregnant women in a rural subdistrict in Narail. The point prevalence of suicidal ideation is 11.4% (95% CI: 8.5% to 15.2%). The prevalence of suicidal thoughts in antenatally depressed women accounts for 10.2% (95% CI: 6.1% to 16.6%) and in non-depressed pregnant women it is 12.2% (95% CI: 8.4% to 17.3%) -according to this study.

The obtained prevalence of suicidal ideation in pregnancy from this study aligns with the meta-analysis conducted by Xiao et. al. involving 23 countries revealing a 10% prevalence of suicidal thoughts in pregnancy [22] but higher than the study performed by Zhang et.al, which unveiled a 6.71% prevalence. The probable reason for this deviation is the selection of the target population; pregnant women only in the third trimester were included in the latter [23].

This study is also in agreement with the global prevalence (2.73%-18%) of suicidal ideation in pregnancy [24] and resembles a similarity with the 11.8% prevalence determined in Ethiopia [25]. 7.6% of women have suicidality in urban India [26]. In the perspective of Bangladesh, 6.5% of adolescent pregnant women attempt suicide [19], whereas 14% of pregnant women at 34-35 weeks of gestational age have thoughts of self-harm [18] which is in line with this study.

Depressive disorder is one of the preeminent causes of suicidal ideation and behavior among Sri Lankan pregnant women, where 7.4% of women develop suicidality during pregnancy and 25% of women have lifetime suicidal ideation [27]. A 3.9% (95% CI:3.0% to 5.1%) suicidality is determined among Brazilian expectant women [28].

Lower-income countries witness 34.0%, (95% Confidence Interval: 33.1%-34.9%) prevalence of prenatal depression which is higher than middle-income countries (22.7%, 95% Confidence Interval: 20.1%-25.2%) [29]. The prevalence of antenatal depression among rural Bangladeshi pregnant women is 33%-39% (95% CI: 33.9% to 44%). Prenatal major depression is revealed as a significant conducive factor to the development of suicidal ideation in some global studies [1, 6]. However, perceived insufficient social support, conceived within the first year of the previous pregnancy, and perceived unfavorable health conditions play crucial roles in generating suicidal thoughts among adolescent expectant mothers in Bangladesh [19]. In this study, the prevalence of suicidal ideation reveals a similarity both in antenatally depressed and non-depressed women (10.2% in depressed and 12.2% in non-depressed pregnant women).

## 5. Limitations

In summary, this study was conducted with a comparatively smaller sample size from a certain subdistrict in Bangladesh due to time and resource limitations. As the data were collected from two hospitals, there might be a chance of a remaining group of women who were unintended to seek medical care without experiencing extreme physical conditions. For that reason, this study might be unable to capture the full diversity of the population leaving a generalizability bias.

## 6. Conclusion

In conclusion, approximately one in every hundred women in Bangladesh bear suicidality during pregnancy and the percentage is almost similar in depressed and non-depressed women. That signifies the role of other contributing factors in the development of suicidal ideation in pregnancy. This study underscores the need to evaluate all contributors to antenatal suicidality to formulate specific plans and programs to minimize the burden of antepartum suicides. In addition, the considerable prevalence of suicidal thoughts among rural Bangladeshi pregnant women indicates the necessity of adequate counseling, care, and support for a woman during her pregnancy.

## Data Availability

Data is available in figshare.com, DOI: 10.6084/m9.figshare.24994110

## Research Data

Data is available on figshare.com. DOI: 10.6084/m9.figshare.24994110

## Acknowledgment

I am profoundly thankful to the Almighty for granting me the opportunity to pursue an MPH (Epidemiology) at North South University. I am also grateful to North South University for allowing me to do the research. I am greatly thankful to Dr. S M Mashud, UH&FPO, Upazila Health Complex, Lohagara, Narail for his unwavering support in the study.

## References

1. Gelaye, B., S. Kajeepeta, and M.A. Williams, Suicidal ideation in pregnancy: an epidemiologic review. Archives of women’s mental health, 2016. 19: p. 741–751.

2. Rao, W.-W., et al., Worldwide prevalence of suicide attempt in pregnant and postpartum women: a meta-analysis of observational studies. Social psychiatry and psychiatric epidemiology, 2021. 56: p. 711–720.

3. Fuhr, D.C., et al., Contribution of suicide and injuries to pregnancy-related mortality in lowincome and middle-income countries: a systematic review and meta-analysis. The Lancet Psychiatry, 2014. 1(3): p. 213–225.

4. Wallace, M.E., et al., Pregnancy-associated homicide and suicide in 37 US states with enhanced pregnancy surveillance. American journal of obstetrics and gynecology, 2016. 215(3): p. 364. e1-364. e10.

5. Lindahl, V., J.L. Pearson, and L. Colpe, Prevalence of suicidality during pregnancy and the postpartum. Archives of women’s Mental Health, 2005. 8: p. 77–87.

6. Newport, D.J., et al., Suicidal ideation in pregnancy: assessment and clinical implications. Archives of women’s mental health, 2007. 10: p. 181–187.

7. World Health Organization, Depression and other common mental disorders: global health estimates. 2017.

8. Kuehner, C., Gender differences in unipolar depression: an update of epidemiological findings and possible explanations. Acta Psychiatrica Scandinavica, 2003. 108(3): p. 163–174.

9. Osman, N.N. and A.I. Bahri, Impact of altered hormonal and neurochemical levels on depression symptoms in women during pregnancy and postpartum period. Journal of Biochemical Technology, 2019. 10(1): p. 16.

10. Dadi, A.F., et al., Global burden of antenatal depression and its association with adverse birth outcomes: an umbrella review. BMC public health, 2020. 20: p. 1–16.

11. Mukherjee, S., et al., Racial/ethnic disparities in antenatal depression in the United States: A systematic review. Maternal and child health journal, 2016. 20: p. 1780–1797.

12. Chatillon, O. and C. Even, La dépression de l’antepartum: prévalence, diagnostic, traitement. L’Encéphale, 2010. 36(6): p. 443–451.

13. Mitchell□Jones, N., et al., Psychological morbidity associated with hyperemesis gravidarum: a systematic review and meta analysis. BJOG: An International Journal of Obstetrics & Gynaecology, 2017. 124(1): p. 20–30.

14. Biaggi, A., et al., Identifying the women at risk of antenatal anxiety and depression: a systematic review. Journal of affective disorders, 2016. 191: p. 62–77.

15. Minamoto, V.B., et al., Dramatic changes in muscle contractile and structural properties after 2 botulinum toxin injections. Muscle & nerve, 2015. 52(4): p. 649–657.

16. Woody, C., et al., A systematic review and meta-regression of the prevalence and incidence of perinatal depression. Journal of affective disorders, 2017. 219: p. 86–92.

17. Mumu, R.T. and D.K. Mitra, Prevalence and associated factors of antenatal depression in rural Bangladesh. medRxiv, 2024: p. 2024.05. 30.24308225.Preprint.

18. Gausia, K., et al., Antenatal depression and suicidal ideation among rural Bangladeshi women: a community-based study. Archives of women’s mental health, 2009. 12: p. 351–358.

19. Li, J., et al., Suicide attempt and its associated factors amongst women who were pregnant as adolescents in Bangladesh: a cross-sectional study. Reproductive health, 2021. 18: p. 1–9.

20. Gavin, A.R., et al., Prevalence and correlates of suicidal ideation during pregnancy. Archives of women’s mental health, 2011. 14: p. 239–246.

21. Yang, W., et al., Relationship between depression and suicidal ideation in pregnant women and its risk factors. Chinese General Practice, 2020. 23(3): p. 305.

22. Xiao, M., et al., Prevalence of suicidal ideation in pregnancy and the postpartum: A systematic review and meta-analysis. Journal of affective disorders, 2022. 296: p. 322–336.

23. Zhang, L., et al., The prevalence of suicide ideation and predictive factors among pregnant women in the third trimester. BMC pregnancy and childbirth, 2022. 22(1): p. 266.

24. Legazpi, P.C.C., et al., Review of suicidal ideation during pregnancy: risk factors, prevalence, assessment instruments and consequences. Psicologia: Reflexão e Crítica, 2022. 35: p. 13.

25. Belete, K., et al., Prevalence and correlates of suicide ideation and attempt among pregnant women attending antenatal care services at public hospitals in southern Ethiopia. Neuropsychiatric disease and treatment, 2021: p. 1517–1529.

26. Supraja, T., et al., Suicidality in early pregnancy among antepartum mothers in urban India. Archives of women’s mental health, 2016. 19: p. 1101–1108.

27. Palfreyman, A., Addressing psychosocial vulnerabilities through antenatal care—depression, suicidal ideation, and behavior: a study among urban Sri Lankan women. Frontiers in psychiatry, 2021. 12: p. 554808.

28. Faisal-Cury, A., et al., Prevalence and associated risk factors of suicidal ideation among Brazilian pregnant women: a population-based study. Frontiers in psychiatry, 2022. 13: p. 779518.

29. Fekadu Dadi, A., E.R. Miller, and L.J.P.o. Mwanri, Antenatal depression and its association with adverse birth outcomes in low and middle-income countries: a systematic review and meta-analysis. 2020. 15(1): p. e0227323.

